# Diagnostic Utility of Low-Pass Whole Genome Sequencing in Prenatal Detection of Chromosomal Abnormalities in an Indian Cohort

**DOI:** 10.1101/2025.07.07.25330717

**Authors:** Akanksha M Tiwari, Kavyashree Padumale, Apoorva Bhandaru, Sushma Gandla, Suparna Maji, Ram Kumar, Honey Sharma, Nithyanandan Thangavel, Sameer Phalke, N Faheena, Shanmukh Katragadda, Vamsi Veeramchaneni, Ramesh Hariharan, Aparna Ganapathy

## Abstract

**Objective:** This study evaluates the diagnostic utility of low-pass whole genome sequencing (LP-WGS) for the detection of chromosomal abnormalities in Amniotic fluid samples (AFS), Chorionic villi samples (CVS) and Product of conception (POC) samples from India.

**Methods:** A total of 1508 prenatal samples including - AFS, CVS and POC were analyzed using LP-WGS at either low-resolution (∼0.5-1X) or high-resolution (∼5X). CNV analysis was performed using StrandNGS v4.2 and Variant Intelligence Applications™ (VIA) v7.0. Copy number variants (CNVs) were classified according to ACMG guidelines with the support from publicly available databases.

**Results:** Among 1508 samples analysed, chromosomal abnormalities were identified in 168 cases resulting in a diagnostic yield of 11.1%. The detection rate was significantly higher in first-trimester samples (44.0%) compared to second-trimester samples (8.2%). Based on clinical indications, abnormalities were detected in 10.1% of cases with ultrasound abnormalities, 4.0% of cases with positive maternal serum screening, and 35.8% of cases flagged as high-risk by non-invasive prenatal testing (NIPT). The commonly assessed ultrasound markers were more prevalent in negative cases than in those with clinical findings highlighting their limited predictive value in isolation.

Aneuploidies accounted for 71% of the detected abnormalities which included 10 mosaic aneuploidies, followed by cytoband-level CNVs (25%). Polyploidy and haploidy were detected in 2.38% of cases, while in 1.6% of the cases gene-level CNVs were detected. CNVs lower than 500kbp, including few gene level CNVs, were detected in the high resolution workflow while the low resolution workflow was effective for events >500Kbp. We detected a mosaic gain of chromosome 13 and chromosome X in a POC sample and mosaic gain of chromosome 21 in an AFS sample which was not detected in QF-PCR due to its technical limitation. The most frequently observed chromosomal abnormality was Down syndrome, followed by Turner syndrome, Edwards syndrome, triple X, trisomy 16, Patau syndrome and 22q11.21 deletion/duplication syndrome. Turner syndrome was the most frequent observation in POC samples. In 12 cases, multiple CNVs were detected, with concurrent gain and loss events suggesting potential underlying balanced translocations in parents, as supported by literature on recurrent cytoband involvement.

Concordance analysis performed on a subset of samples showed a 100% concordance with whole exome sequencing (WES) and a partially reduced concordance of 98% with QF-PCR possibly due to lower sensitivity of detecting mosaic events by QF-PCR. Unlike QF-PCR, LP-WGS test could detect mosaic CNV, cytoband-level CNVs and trisomies involving chromosomes not routinely targeted by the assay, such as trisomy 19, 16, 10, 17, 4, 5, 2, frequently associated with first trimester miscarriages.

**Conclusions:** LP-WGS is a robust, accurate, cost effective tool for prenatal detection of chromosomal abnormality. It offers a high diagnostic yield with rapid turnaround time, and the ability to detect a broad spectrum of abnormalities. It has the potential to detect chromosomal anomalies associated with first trimester miscarriage, mosaic copy number variants and possible unbalanced translocation events. These findings support its utility as a first-line method in prenatal diagnostics.

## INTRODUCTION

Prenatal genetic testing is a crucial aspect of modern antenatal care, providing expectant parents with vital information regarding potential genetic disorders in the developing fetus ^1^. It is particularly recommended in pregnancies with elevated risk such as those involving advanced maternal age, family history of genetic disorders or structural abnormalities detected via ultrasound. This testing encompasses both screening and diagnostic methods, allowing for the assessment of various genetic conditions, including aneuploidies and inherited disorders.

Common screening tests include Noninvasive Prenatal Testing (NIPT), which detects common aneuploidies from the cell-free fetal DNA in maternal blood ^1 2^. Invasive diagnostic tests such as chorionic villus sampling (CVS) and amniocentesis provide definitive genetic diagnoses but carry a risk of complications ^3^.

Common fetal genetic causes of congenital abnormalities include several chromosomal disorders like trisomy 21 or Down syndrome, Turner syndrome and Klinefelter syndrome. Additionally, microdeletion syndromes, caused by small chromosomal deletions, can lead to a range of abnormalities ^4^. Advances in genomic technologies have enabled the identification of various genetic causes, including chromosomal disorders, copy number variants (CNVs), and monogenic disorders. Early detection of these abnormalities is essential for optimizing management and outcomes.

Globally, birth defects occur in about 3% to 5% of all live births and are significant contributors to infant morbidity and mortality ^5 6^. They are often linked to genetic causes, including chromosomal abnormalities and specific genetic mutations that contribute to 20–25% of congenital malformations ^7^. In India, the prevalence of birth defects is estimated at 0.84% to 2.9% ^8 9^. These anomalies account for 8-15% of perinatal deaths and 13-16% of neonatal deaths ^10^.

Verma et. al., 2018 reports that the performance of SNP-based NIPT in India is comparable to global standards, demonstrating its feasibility and reliability in the Indian population despite genetic diversity ^8^. This suggests that NIPT could be a valuable screening tool in the Indian prenatal setting. Furthermore, systematic reviews and case studies on fetuses with increased nuchal translucency or cystic hygroma emphasize the importance of comprehensive genetic testing, including karyotyping, CMA, and multigene panels, to improve diagnostic yields ^11^. Overall, these studies collectively demonstrate that advanced genomic techniques are increasingly integral to prenatal diagnosis in India, with a trend towards higher diagnostic yields and more precise etiological understanding. However, challenges such as limited access to molecular testing and the need for specialized infrastructure remain significant barriers. The integration of these technologies into routine prenatal care holds promise for improved outcomes, but requires concerted efforts to expand availability and expertise across the country.

Low-pass whole genome sequencing (LP-WGS) is emerging as a cost-effective and reliable alternative for detecting copy number variants (CNVs) in clinical settings, particularly in low- and middle-income countries ^12^. Conventional cytogenetic and molecular techniques such as karyotyping, chromosomal microarray analysis (CMA), and quantitative fluorescent PCR (QF-PCR) have long served as the cornerstone of prenatal genetic diagnostics. While karyotyping facilitates detection of large chromosomal abnormalities, its resolution is limited. CMA enhances detection of submicroscopic CNVs, yet it requires higher costs and technical complexity ^13^. QF-PCR is targeted, rapid, and cost-effective, but lacks genome-wide coverage and is constrained to specific abnormalities.

LP-WGS is rapidly emerging as a transformative and cost-effective approach for detecting CNVs and structural variations (SVs), particularly in prenatal genetic testing. By utilizing low-depth sequencing combined with robust bioinformatics algorithms, LP-WGS provides genome-wide coverage and enables reliable detection of chromosomal abnormalities at a resolution comparable to that of CMA, while significantly reducing cost and turnaround time ^14 15 16^. This method offers diagnostic yields similar to traditional CMA but with lower technical complexity and greater scalability ^16^. Its adaptability to both prenatal and postnatal settings, coupled with its affordability, makes LP-WGS particularly well-suited for clinical implementation in resource-limited environments such as India. Moreover, LP-WGS has shown the potential to standardize CNV detection across diverse platforms, reduce dependence on multiple assays, and streamline workflows representing a key advancement toward equitable access to genomic medicine in prenatal care.

Low-resolution LP-WGS demonstrates a significant capability in identifying CNVs that exceed 500 kilobases in size, exhibiting remarkable sensitivity^17^, whereas High-Resolution LP-WGS can detect CNVs over 50 kilobases with the ability to detect low-level mosaicism and increase diagnostic yield by detecting a wider range of variants, including smaller CNVs, intragenic CNVs and rare variants^18^.

Ultrasound soft markers and structural anomalies detected through ultrasound are frequently linked to chromosomal irregularities ^19^, suggesting an increased probability of genetic issues. The rate at which chromosomal abnormalities are identified is notably higher in fetuses with ultrasound-detected anomalies compared to those without such findings. In such cases, the use of genome-wide technologies like LP-WGS enhances the likelihood of identifying a genetic cause.

In this study, we present a comprehensive evaluation of LP-WGS as a first tier test for prenatal diagnosis in a clinical setting. Using our validated CNV detection pipeline, we analyzed over 1500 prenatal samples referred for chromosomal analysis due to abnormal ultrasound findings or positive screening results. To our knowledge, this is the largest dataset from the Indian population assessing the utility of LP-WGS for the detection of chromosomal aneuploidies and clinically relevant copy number variations in a prenatal cohort. Through this work, we aim to demonstrate the diagnostic yield, technical performance and practical feasibility of implementing LP-WGS in routine prenatal care. Our findings highlight the potential of this approach to provide cost effective, genome-wide insights in a scalable manner, addressing a critical need for accessible genomic diagnostics in India.

## METHODS

### Patients

This study includes 1,508 fetuses with structural or growth anomalies, increased anuploidy risk identified by ultrasound, or positive prenatal screening test results. Samples were referred to our laboratory from various hospitals/clinics from across India, between December 2023 and May 2025. Informed consent was obtained from all subjects and sequencing of patient samples was approved by the Institutional Ethics Committee of Strand Life Sciences.

### Sample preparation

Genomic DNA was extracted from amniotic fluid (AFS), chorionic villus sampling (CVS), products of conception (POC), fetal and peripheral blood using a genomic DNA extraction kit (Cambrian Bioworks, LLP) according to the manufacturer’s instructions. For amniotic fluid, CVS and POC samples, an additional Maternal Cell Contamination (MCC) test was performed using STR analysis to detect the presence of maternal DNA contamination in the fetal sample. Fetal samples with >15% maternal contamination did not qualify for further genetic testing. Samples showing maternal cell contamination were subjected to cell culture to enrich the fetal cells and to reduce or eliminate maternal DNA contamination before proceeding with prenatal genetic analysis.

### Library preparation

The extracted DNA underwent whole genome library preparation using xGen™ DNA Library Prep EZ Kit (Integrated DNA Technologies, Inc) as per manufacturer’s instructions. Approximately 25 ng of input gDNA was used for the process, which included enzymatic fragmentation, end repair and adapter ligation. A 0.8X purification was subsequently performed to isolate adapter-ligated fragments. These purified fragments were then barcoded through limited cycles of PCR amplification to generate the whole genome library. The quantity and quality of the library was assessed using the Qubit™ dsDNA HS Assay Kit (ThermoFisher Scientific, USA) and the Tapestation 4200 (Agilent Technologies, USA). The quantified libraries were normalized, pooled and sequenced on the Novaseq 6000 or Novaseq X Plus platform (Illumina) using 150 bp paired-end chemistry, following the manufacturer’s instructions.

### NGS based Copy Number Analysis

The copy number variant (CNV) analyses methods are categorized as Low Resolution (LR) and High Resolution (HR) based on the detection threshold: ≥500 Kb for LR and ≥50 Kb for HR. The sequenced reads were aligned to the human genome (hg19) using the Illumina DRAGEN germline pipeline v4.1.23 to generate BAM files. CNV analysis was performed using two platforms: Variant Intelligence Applications™ (VIA) v7.0 (https://bionano.com/via-software/) and StrandNGS v4.2 (http://www.strand-ngs.com/). Additionally a coverage based independent method was employed to determine the ploidy for each chromosome, and when available, the findings were verified with QF-PCR for both common aneuploidies, and polyploidies in the sample. This approach uses read counts for each chromosome from healthy (diploid) reference samples and compares the ratios to those in the test sample for each chromosome. Details of the NGS based Copy Number Analysis are provided in the online supplementary resource.

### Copy Number Variant interpretation

Interpretation and analysis of CNVs was done with the help of databases such as Clinical Genome Resource (ClinGen), Online Mendelian Inheritance in Man (OMIM), DECIPHER grch37, UCSC Genome Browser hg37, Database of Genomic Variants (DGV) and ClinVar (June 2023). CNVs were assessed for pathogenicity of germline duplications and deletions following the American College of Medical Genetics and Genomics (ACMG) guidelines ^20^ and were classified into five categories: 1) pathogenic, 2) likely pathogenic, 3) variant of uncertain significance (VUS), 4) likely benign, and 5) benign. Primarily, likely pathogenic (LP) and pathogenic (P) variants were considered for reporting, however, a few variants of uncertain significance (VUS) were also considered for reporting.

### Secondary Data Validation

Among the referred cases, a set of samples were referred for parallel testing either with whole exome sequencing (WES) or QF-PCR. These cases were retrospectively analyzed for validation and concordance analysis for LP-WGS. WES findings were available for 41 out of 1,508 samples.

QF-PCR data was available for 1,421 out of 1,508 samples. DNA isolated from Amniotic Fluid/CVS/POC underwent multiplex PCR amplification using Quantitative Fluorescent Polymerase Chain Reaction (QF-PCR) technology using a commercial kit according to manufacturer’s instructions. The size of the DNA fragments and their corresponding fluorescent signals were analyzed and interpreted using GeneMapper™ Software to detect common aneuploidies.

## RESULTS

### Prenatal sample characteristics and reason for referral

A total of 1511 prenatal samples were received for LP-WGS analysis during the period Dec 2023 to May 2025. A failure rate of 0.19% (3/1511) was noted in this cohort. Of these, 1508 samples were considered for further processing. 1109 samples were processed using low-resolution assay and 399 samples underwent high-resolution analysis (Fig 1A). The prenatal specimen type includes AFS, CVS, and POC with AFS representing the largest proportion of the cohort, around 1,255 (83.2%) samples, primarily collected during the second trimester. POC samples obtained primarily following pregnancy loss constituted 160 (10.6%) of the total samples received. CVS samples made up the remainder of the cohort with 93 (6.2%) CVS samples collected during the first trimester (Fig 1B). Failure rate observed in our cohort was around 0.19%

**Figure 1.**
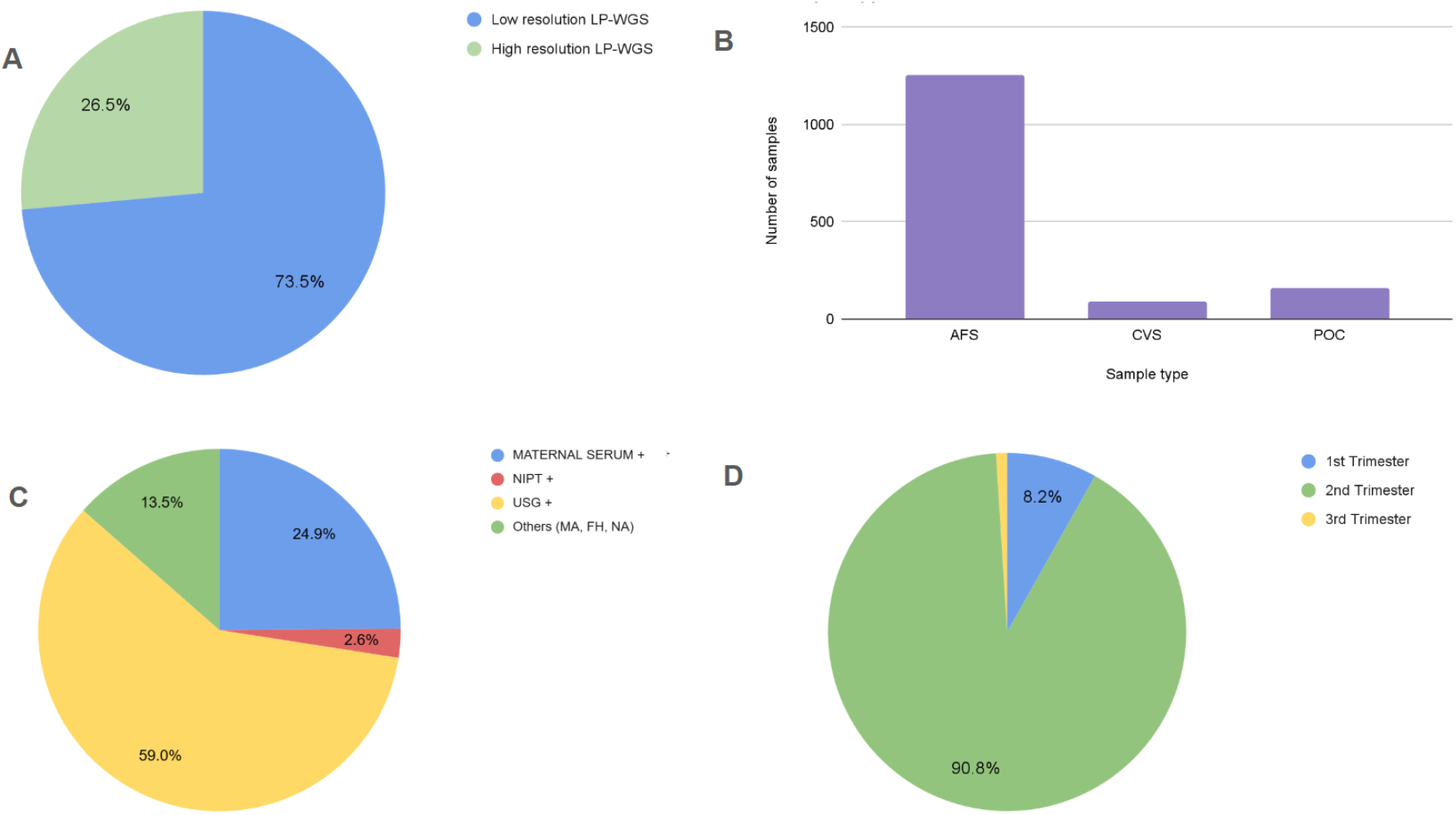
Overview of LP-WGS cases analyzed: Distribution of test type, sample type, referral reasons and trimester. A. The pie chart shows the distribution of 1,508 samples, with 1,109 (73.6%) analyzed using low-resolution LP-WGS and 399 (26.4%) processed using high-resolution LP-WGS. B. Distribution of prenatal sample types in the cohort: Amniotic fluid samples (AFS) comprised the majority 1,238 (82%), followed by product of conception (POC) 164 (10.87%) and Chorionic villus samples (CVS) 91 (6.03%). C. Clinical indications for referral: Ultrasound abnormalities were the most common indication in 889 cases followed by elevated risk based on double, triple, or quadruple marker tests in 375 cases, and 39 cases flagged high-risk through NIPT. Other reasons for referral included - MA (missed abortion), FH (family history) and NA (detail not available). D. Trimester wise sample distribution: The majority (∼90%) were received during the second trimester of pregnancy, while 8.2% were obtained in the first trimester and 1% in the third trimester.

The main reasons for referral included abnormal ultrasound findings, intermediate or high risk from maternal serum screening results or NIPT, adverse obstetric history and relevant family history. Among the 1,508 cases received, approximately 890 had abnormal ultrasound findings, 375 were referred based on elevated risk from maternal serum screening (double, triple, or quadruple marker tests), and 39 were flagged as high-risk through NIPT (Fig 1C). Nearly 90% of the samples were received during the second trimester of pregnancy, while only 8% were obtained in the first trimester (Fig 1D).

### Diagnostic yield of LP-WGS in prenatal diagnosis

In our cohort of 1,508 prenatal samples analyzed using LP-WGS, chromosomal abnormalities were identified in 168 cases, yielding an overall detection rate of 11.1% (Fig 2A). Among these, 136 positive cases were analyzed using low-resolution workflow and 32 using high resolution workflow yielding a detection rate of 12.3% and 8% respectively (Fig S1). The detection rate was notably higher in first trimester miscarriage and IUD samples 57/160 (35.6%) compared to samples from ongoing pregnancies with live fetuses 111/1348 (8.2%).

**Figure 2.**
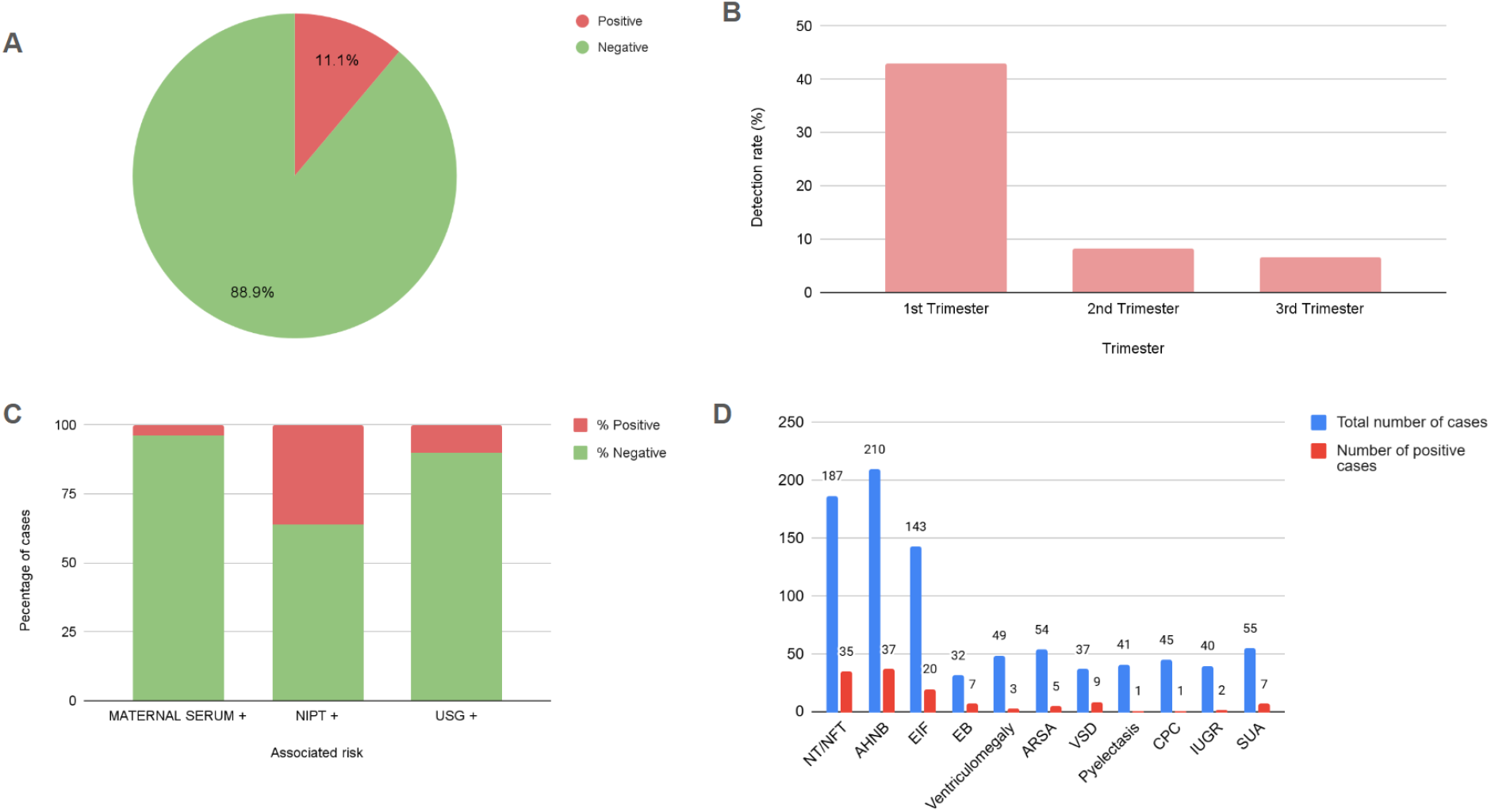
Diagnostic yield and USG soft marker distribution among LP-WGS prenatal cases. A. Pie chart showing the overall detection rate of chromosomal abnormalities in the cohort (n = 1,508) analyzed using LP-WGS. Abnormalities were detected in 168 cases, corresponding to a detection rate of approximately 11.1%. B. Trimester-wise distribution of cases and detection rates: In the first trimester, chromosomal abnormalities were detected in 55 out of 125 cases (44.0%), while in the second trimester, abnormalities were identified in 112 out of 1,366 cases (8.2%). C. Distribution of chromosomal abnormalities based on clinical indications: Abnormalities were detected in 90 cases with ultrasound abnormalities (10.1%), 15 cases with positive maternal serum screening (4.0%), and 14 cases flagged as high-risk by NIPT (35.8%). D. Prevalence of common soft ultrasound markers in cases with and without clinically significant findings: The most commonly observed soft ultrasound markers were nuchal translucency (NT)/nuchal fold thickness (NFT), absent or hypoplastic nasal bone (AHNB), and echogenic intracardiac focus (EIF). NT/NFT was seen in 35 of positive cases out of 187 cases; AHNB in 37 out of 210 cases and EIF in 20 out of 143 total cases where these soft markers were observed.

We also assessed diagnostic yield based on common reasons for referral such as USG abnormality (with or without other screening tests) and high risk in maternal serum screening and NIPT (with or without other screening tests). Chromosomal abnormalities were detected in 91 cases with ultrasound abnormality (10.2%), 15 of the maternal serum screening positive cases (4.0%), and 14 of the NIPT-positive cases (35.8%) (Fig 2C). NIPT-positive cases were associated with greater risk of underlying chromosomal aberration, highlighting its better predictive value compared to other screening tests. We also assessed the number of cases received and detection rate in each trimester, among 123 first-trimester samples, abnormalities were detected in 53 cases, corresponding to a detection rate of 43%. It is interesting to note that 85 samples received in the first trimester were POC samples and in 45 cases (52%) we had a positive finding which indicates that early pregnancy loss is frequently associated with chromosomal anomalies. In comparison, 1,368 samples (1301 fetus and 67 POC samples) were received during the second trimester, with chromosomal abnormalities identified in 114 cases, resulting in a detection rate of 8.3% (Fig 2B).

### Prevalence of ultrasound soft markers

We evaluated the prevalence of common USG soft markers such as nuchal translucency (NT)/nuchal fold thickness (NFT), absent or hypoplastic nasal bone (AHNB), and echogenic intracardiac focus (EIF) in cases with positive findings. NT/NFT was observed in 35 positive cases out of 187 total cases with this soft marker, AHNB was seen in 37 positive cases out of 210 cases, while EIF was present in 20 positive cases out of 143 cases with this soft marker (Fig 2D, Table S2). We evaluated common USG findings in cases of Down syndrome, Edwards syndrome, Patau syndrome, Turner syndrome, Triple X, Klinefelter syndrome and all other microdeletions and microduplications; this excluded all POC cases. Among 54 cases of Down syndrome, the most common USG soft marker was AHNB (22) followed by increased NT (17), EIF (15), VSD (Ventricular septal defect) (6), ARSA (Aberrant Right Subclavian Artery) (4), EB (Echogenic Bowel) (4) and ventriculomegaly (2). Other common USG findings included Tricuspid regurgitation, ductus venosus agenesis, AVSD and increased prenasal thickness (Table S3 ^21–24^).

### Prenatal detection of chromosomal abnormalities

Among 168 cases with pathogenic/likely pathogenic findings, aneuploidies constituted the majority (71%), followed by cytoband-level CNVs (25%). Polyploidy and haploidy constituted 2.38% of cases which were mainly POC samples, while in 1.6% of the cases, smaller gene-level CNVs were detected (Fig 3A). Most frequently observed syndrome in prenatal samples was trisomy 21 (Down syndrome), followed by Turner syndrome, Edwards syndrome, triple X, trisomy 16, Patau syndrome, Klinefelter syndrome, trisomy 10 and 22q11.21 deletion/duplication syndrome. We received around 160 POC cases of which 57 cases had a positive finding with Turner syndrome detected most frequently, followed by Down syndrome, Edwards syndrome, trisomy 16, triploidy, 22q duplication syndrome, and 15q duplication syndrome (Figure 3B and 3C). Interestingly, majority of the POC cases with positive findings (45 out of 57) were those with spontaneous abortion in the first trimester and in these cases, in addition to commonly observed aneuploidies, we detected trisomy 16, 10, 17, 5, 4, and 2, highlighting that these chromosomal anomalies are largely incompatible with early fetal survival.

**Figure 3.**
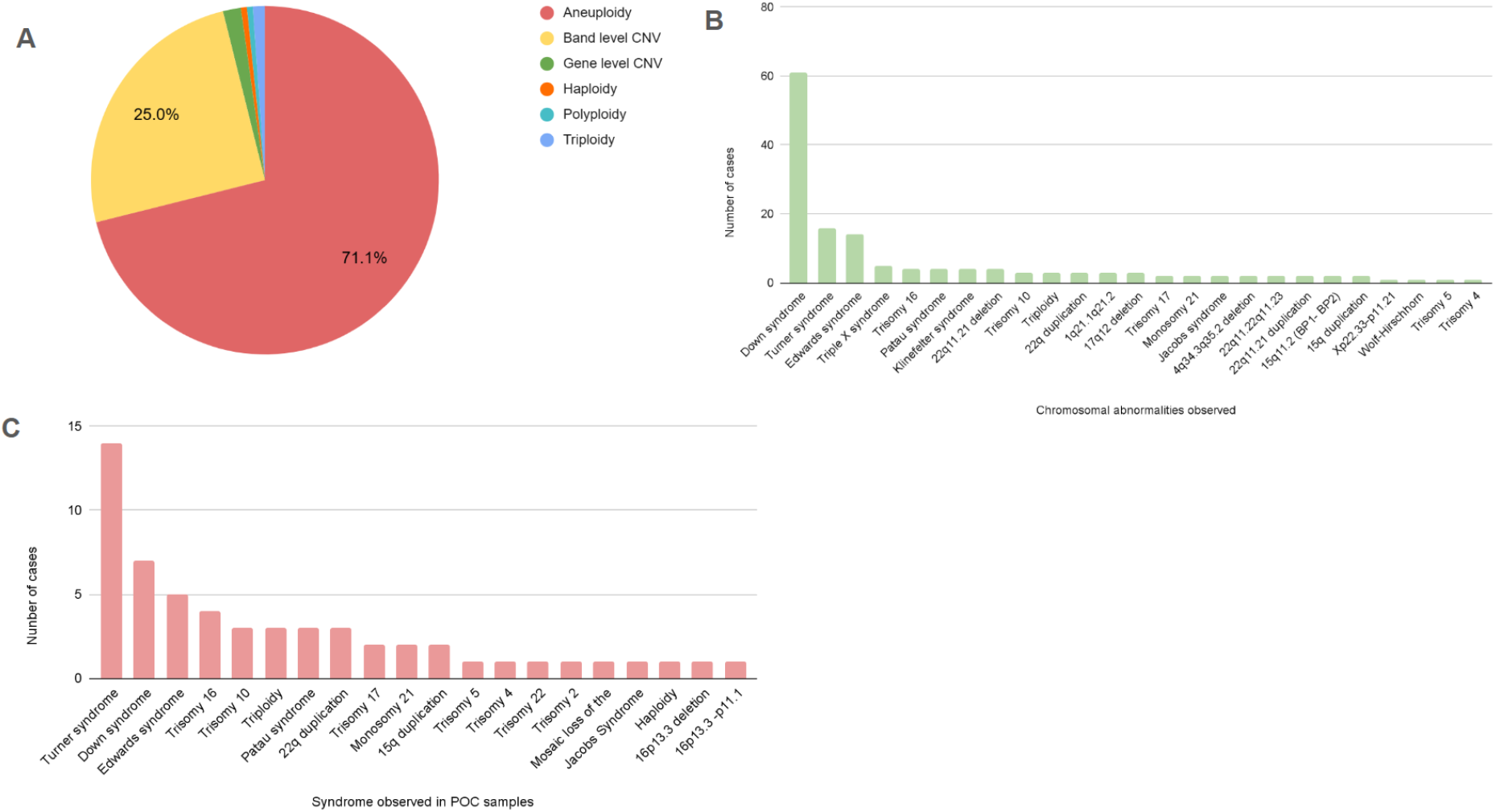
A. The pie chart illustrates the distribution of detected chromosomal abnormalities, with aneuploidies detected in majority of the cases (71%), followed by cytoband-level CNVs (25%). Polyploidy, haploidy, and gene-level CNVs accounted for the remaining proportion. B. Among the 1508 prenatal cases in the cohort, Down syndrome was the most commonly reported aneuploidy, observed in 61 cases. This was followed by Turner syndrome, Edwards syndrome, and triple X syndrome. C. Out of 160 cases of missed abortions, 57 (35.6%) exhibited chromosomal abnormalities. Turner syndrome was the most frequently detected abnormality, followed by Down syndrome, Edwards syndrome, trisomy 16, triploidy, 22q11.2 duplication syndrome, and 15q duplication syndrome.

136 pathogenic and likely pathogenic CNVs were detected in low-resolution cases, of which the majority (107 CNVs) were larger than 40 Mb (upto 243 Mbp) and constituted all common and rare aneuploidies. 15 CNVs ranged between 10–40 Mb in size, 18 between 1–10 Mb, and 4 CNVs were sized between 500 kb and 1 Mb. In comparison, among the 32 pathogenic/likely pathogenic CNVs detected in high-resolution cases, 19 were larger than 40 Mb, 4 ranged from 10–40 Mb, 5 from 1–10 Mb, 2 CNVs between size 500 Kb and 1 Mb and 4 CNVs were smaller than 500 kb, which fall below the detection limit of low resolution test. These findings highlight that while both platforms predominantly identified large CNVs, the high resolution assay offered improved sensitivity for detecting smaller CNVs, particularly those under 500 kb (Figure S3).

We reported 175 pathogenic or likely pathogenic variants and 5 variants of uncertain significance (VUS) in 168 positive cases. In accordance with ACMG and ACOG guidelines for prenatal testing ^25,26^, only pathogenic and likely pathogenic variants were routinely considered for reporting. However, variants of uncertain significance (VUS) were reported in select cases where the CNV size exceeded 1 Mb, especially when limited published evidence suggested potential clinical relevance, or when reporting was specifically requested by the referring clinician.

In 12 positive cases (6.7%), we identified multiple CNVs with concurrent copy number gains and losses within the same sample. Interestingly, the affected cytobands Xp22.33, Xq25, 7q36, 6p22, 11q23.2, 16p13.3, 14q11.1-q21.3, and 9p24.3-p23 have each been independently reported in the literature in association with chromosomal translocation events. These findings raise the possibility of underlying balanced translocations in one of the parents, which may predispose to unbalanced rearrangements in the fetus. While LP-WGS cannot detect balanced translocations, it can reveal the resulting unbalanced rearrangements as independent copy number gains and losses (Table S1).

### Concordance with WES and QF-PCR-

We also compared the results from cases where WES (41 cases) and QF-PCR (1,421 cases) were performed in parallel. The overall concordance between LP-WGS and WES was 100% in regards to CNVs. In addition to SNVs WES can detect all large CNVs that can be identified using LP-WGS, the latter has greater relevance in prenatal settings due to low-cost and rapid turnaround time. Among the LP-WGS negative cases, a confirmatory diagnosis was established in 8 cases based on pathogenic or likely pathogenic SNVs identified (Table S4). For QF-PCR the concordance with LP-WGS was 98.07%, based on the analysis of common aneuploidies detectable by QF-PCR, specifically chromosomes 13, 18, 21, and gonosomal aneuploidies. Discordance between LP-WGS and QF-PCR was due to the mosaic nature of the chromosomal aneuploidies, which QF-PCR failed to detect. Additionally, QF-PCR is limited in its ability to identify cytoband-level CNVs and trisomies involving chromosomes not routinely targeted by the assay, such as trisomy 19, trisomy 16, 10, 17, 4,5,2 which were detected mainly in first trimester spontaneous miscarriages. These findings further support the reliability and broader diagnostic utility of LP-WGS in prenatal testing.

## DISCUSSION

This study was aimed to evaluate the clinical utility of LP-WGS for detecting chromosomal abnormalities in a large prenatal cohort. The primary aim was to determine whether LP-WGS could serve as a reliable first-tier diagnostic tool across diverse clinical indications and specimen types in the prenatal setting. Specifically, we assessed detection rates across different trimesters, the significance of screening markers, and concordance with QF-PCR and WES, hypothesizing that LP-WGS would provide a robust detection rate for pathogenic CNVs and aneuploidies, including those often missed by conventional cytogenetic methods, while offering advantages in turnaround time, resolution, and scalability.

We analyzed 1,508 prenatal samples over 18 months using LP-WGS, which showed an overall diagnostic yield of 11.1% (Fig 2A). Interestingly, the low-resolution workflow yielded a higher detection rate 12.3% than the high-resolution workflow 8% which aligns with previously reported detection rates from prenatal LP-WGS and chromosomal microarray studies ^23-24^ (Fig S1). This difference likely reflects referral patterns: cases suspected to have larger, chromosome-level abnormalities, such as those with abnormal ultrasound findings or prior aneuploidy risk were more often routed to the low resolution workflow. In contrast, referrals for the high resolution workflow were relatively fewer and typically not indicated when common aneuploidies were already suggested by screening tests like serum markers or NIPT. This inherent selection bias is likely to contribute to the observed difference in detection rates between the two workflows.

Notably, the highest detection rate was observed in first-trimester samples 43%, compared to 8.3% in the second trimester, highlighting the potential of early gestation testing for timely and actionable insights (Fig 2B). This is consistent with the known higher prevalence of chromosomal abnormalities, particularly aneuploidies such as trisomy 21 and Turner syndrome in early pregnancy losses and the first trimester, often due to embryonic lethality associated with severe chromosomal anomalies ^34 35^. The majority of the samples (82%) were derived from amniotic fluid, reflecting the widespread clinical adoption of second-trimester amniocentesis. In contrast, only 6.2% of cases involved CVS, indicating a continued underutilization of first-trimester invasive diagnostics, despite evidence suggesting higher diagnostic yields during this window ^27-28^ (Fig 1B).

In our cohort, ∼15 CVS samples in early gestation were referred for NT, HNB, abnormal serum markers, or structural anomalies. Among these, chromosomal abnormalities were confirmed in six cases of Down syndrome along with other clinically significant findings highlighting the value of early testing when such markers are present. POC made up 10.6% of the cohort, primarily submitted following pregnancy loss (Fig 1B). Most referrals followed ultrasound or serum screening, with fewer from high-risk NIPT. Although first-trimester samples had higher yield, most testing occurred in the second trimester, possibly reflecting hesitancy toward early procedures like CVS despite their diagnostic advantage ^37^. The varied clinical indications and sample types underscores the versatility of LP-WGS across different diverse prenatal contexts.

Among clinical indications, NIPT-positive referrals exhibited the highest diagnostic yield, 35.8%, followed by ultrasound abnormalities, 10.2%, and serum screening-positive cases, 4.0%, reaffirming the predictive value of non-invasive screening and targeted imaging ^38^ (Fig 2C). NIPT poses no risk of miscarriage and has good positive predictive value, making it an attractive early screening option. Traditionally, ultrasound markers such as increased NT, AHNB, and EIF have been considered indicative of underlying chromosomal abnormalities. However, our findings suggest that this assumption may not always hold true. For instance, NT/NFT was observed in 2.32% of positive cases compared to 10.08% of negative cases, hypoplastic NB was seen in 2.45% of positive and 11.47% of negative cases, while EIF was present in 1.33% of positive cases and 8.16% of negative cases (Fig 2D, Table S2). These markers were more frequently present in chromosomally normal cases than in abnormal ones. These findings are consistent with previous studies showing that isolated soft markers, especially EIF and hypoplastic nasal bone, are frequently seen in fetuses with normal karyotypes highlighting the need for cautious interpretation of these markers in prenatal screening ^39^ However, when NT and NB occurred concurrently, as seen in 11 of 27 such cases, the diagnostic yield significantly increased with Down syndrome detected in 9 of these cases.

The spectrum of pathogenic findings was dominated by aneuploidies, 71%, followed by band-level CNVs, 25%, and a small proportion of polyploidies and gene-level events (Fig 3A). Concordance analysis with 41 WES and 1,421 QF-PCR cases highlighted the superior diagnostic performance of LP-WGS. It showed 100% concordance with WES confirming its reliability for CNV detection and 98.07% concordance with QF-PCR, primarily due to QF-PCR’s limitations in detecting mosaicism (Table S1). Notably, all CNVs that were concordant between LP-WGS and WES were large events, which both platforms are capable of detecting. While WES, in principle, can detect all CNVs identifiable by LP-WGS, the latter offers significant practical advantages in the prenatal setting. Given its faster turnaround time and robust detection of chromosomal anomalies, LP-WGS remains a strong candidate for a first-tier test when such abnormalities are suspected and clinical decision-making is time-sensitive.

Among the 1,508 prenatal samples, aneuploidies constituted 67% of the abnormalities, with Down syndrome most common (Fig 3B). Out of 61 Down syndrome cases, increased NT, hypoplastic nasal bone and EIF were the most frequent soft markers consistent with prior studies emphasizing NT and NB as key first-trimester indicators for trisomy 21^40 41^. In missed abortion cases, chromosomal abnormalities were found in ∼35.6% (57/160) of samples, aligning with reports that over 50% of first-trimester losses involve chromosomal defects. ^42^. Turner syndrome was most frequent in POC samples (57/160) highlighting the link between monosomy X and early miscarriage, consistent with the literature ^43 44^ followed by Down and Edwards syndrome, trisomy 16, triploidy and recurrent microduplications such as 22q and 15q (Fig 3B and 3C). Other detected trisomies like 10, 17, 5, 4, and 2 reinforce the impact of severe chromosomal imbalance in early pregnancy loss. Microdeletion/duplication syndromes, often undetectable by conventional cytogenetics were also identified, underscoring LP-WGS’s added diagnostic value. Among the 179 positive cases, 6 mosaic events were detected exclusively by LP-WGS reflecting its superior sensitivity for mosaicism ≥30%, consistent with previous reports ^45,46^. 6.7% of positive cases had multiple CNVs within a single sample, in regions associated with rearrangements and translocation hotspots. While LP-WGS cannot detect balanced translocations, such CNVs may suggest structural complexity, warranting further investigation by karyotyping or FISH (Table S1).

Our results also provide insights into clinical classification of detected variants, with pathogenic or likely pathogenic variants identified in 97.2% of positive cases. Only five VUS were reported, reflecting a conservative strategy and high interpretive confidence. VUSs were considered for reporting only when the clinical findings were unclear and potentially relevant, in line with best practices to reduce patient anxiety while ensuring diagnostic clarity. The VUSs were primarily classified as uncertain due to limited clinical correlation or insufficient literature evidence. Reporting was considered when VUS exceeded 1 Mb, matched the phenotype, or upon clinician request. This highlights the evolving challenges of variant interpretation in prenatal diagnostics. While the prevalence of VUS varies across studies, ^47 48^ prior research underscores both their potential clinical value and the importance of cautious interpretation to prevent unnecessary parental anxiety, consistent with current guidelines ^49 50 51^

In prenatal diagnostics, certain CNVs were excluded from reporting due to limited clinical relevance, high population frequency, uncertain penetrance, or presence in technically challenging genomic regions prone to artifacts. For instance, deletions in exons 1–3 of the *KANSL1* gene are omitted due to complex structure of the 17q21.31 locus, frequent benign polymorphisms, and lack of association with Koolen-de Vries syndrome ^52 53^ Similarly, CNVs at 16p13.11 ^54 55 56^, 15q13.1–q13.3 ^57 58^ and 15q11.2 BP1–BP2 ^59 60^ are often inherited, lack consistent prenatal phenotypes, and show variable or incomplete penetrance with studies showing many postnatal presentations being benign. Gene-level CNVs in complex regions were typically excluded from LP-WGS reporting due to technical constraints, with targeted assays when clinically indicated. These exclusions follow ACMG guidelines to ensure that prenatal reports remain actionable, reduce parental anxiety, and support evidence-based counseling ^26 61^

Our prenatal cohort of 1,508 samples highlights the clear advantages of LP-WGS in routine prenatal diagnostics. In line with previous studies, LP-WGS provided unbiased, genome-wide CNV detection, unconstrained by probe selection as in CMA. In our dataset, LP-WGS identified not only common aneuploidies and large CNVs (>40 Mb), but also clinically significant smaller variants (≤500 kb) and mosaic events that are often missed by targeted methods. Prior validation studies support this- For example ^62^ prior studies reported that LP-WGS identified 17 additional clinically significant CNVs in a cohort of 1,023 cases undetected by CMA, with 98–100% concordance. Similarly, in a cohort of 409 samples, LP-WGS achieved 100% sensitivity for all CNVs previously detected by CMA and additionally uncovered pathogenic CNVs that CMA failed to identify ^63^. LP-WGS’s low DNA input requirement (25–150 ng) and rapid turnaround time proved especially useful for POC and CVS samples, aligning with published data on its utility in low-yield or urgent prenatal scenarios ^64 17 64^.

Despite its robust diagnostic performance, LP-WGS has several inherent limitations that must be considered. It cannot detect balanced rearrangements (e.g., translocations, inversions), polyploidies beyond triploidy (69,XXY), loss of heterozygosity, uniparental disomy (UPD), single-gene disorder and point mutations, While mosaic events ≥30% were reported, low-level mosaicism (> 10%) can also be detected by LP-WGS. However, reporting low level mosaicism requires cautious reporting due to technical limitations in detection accuracy and the risk of false positives, challenges from distinguishing confined placental mosaicism from true fetal involvement and possibility of variable outcomes. LP-WGS also lacks resolution in complex, repetitive regions such as those involving *SMN1, HBA1/2*, due to mapping ambiguity ^65 65^. When clinically indicated, findings in these regions require secondary methods. Moreover, LP-WGS cannot assess the short arms (p-arms) of acrocentric chromosomes (13, 14, 15, 21, 22) due to their repetitive, non-coding structure. However, the absence of disease-associated genes in the p-arm of these chromosomes confines the clinical significance to only their q-arm.

Our findings support LP-WGS as a first-line diagnostic tool, enabled by standardization parameters such as optimal read depth (∼30 million reads), CNV-calling pipelines, and uniform reporting practices. In conclusion, our study highlights LP-WGS as a robust, scalable platform for the detection of chromosomal abnormalities in prenatal settings. With strong diagnostic performance and concordance across methods, it shows promise as a first-tier test supporting timely and informed reproductive decision-making. Though limitations exist, these gaps can increasingly be bridged through integration with complementary technologies and evolving bioinformatics tools. Importantly, the continual expansion of curated genomic databases like DECIPHER ^66^, ClinGen ^67^ and DGV ^68^ along with rising volumes of population and patient-level sequencing data, is enhancing CNV interpretation, reducing uncertainty and supporting evidence-based reporting. As NGS platforms improve in resolution and sensitivity, and as patient-derived variant data continues to grow, these advances will collectively address current limitations and refine the diagnostic landscape. Thus, LP-WGS, when combined with robust datasets and orthogonal validation, offers an increasingly comprehensive solution for prenatal genome analysis.

## Supporting information

https://drive.google.com/drive/u/0/folders/17UdFCTHFq65zaKJF0IylR7Tn04eNojMp

## Data Availability

All data produced in the present work are contained in the manuscript

## Acknowledgements

We thank the patients and families, who consented to participate in this study. We thank all the physicians, who referred the patients to our centre. We also thank the Strand Life Sciences laboratory, bioinformatics, interpretation and genetic counselling staff for providing the infrastructure needed for this study.

