## Supplementary material for "Diagnostic Utility of Low-Pass Whole Genome Sequencing in Prenatal Detection of Chromosomal Abnormalities in an Indian Cohort": https://drive.google.com/drive/u/0/folders/17UdFCTHFq65zaKJF0IylR7Tn04eNojMp

**Supplementary Table S1: List of chromosomal abnormalities identified in this study.**

| S.No. | Sample ID | LR/HR | Trimester (1st, 2nd, 3rd) | Risk | Chromosome Region | ISCN Nomenclature | Length (Kb) | Classification | Syndrome Name | QF-PCR Result | WES Result |
|---|---|---|---|---|---|---|---|---|---|---|---|
| 1 | S-0009 | LR | 1st | NA | chr17:1-81,195,210 | (17)x3 | 81195.2 | Pathogenic | Trisomy 17 | NP | NP |
| 2 | S-0012 | LR | 1st | USG+ | chrX:1-155,270,560 | Xp22.33q28(1_154760280)x1 | 154760.3 | Pathogenic | Turner syndrome | Concordant | NP |
| 3 | S-0020 | LR | 2nd | USG+ | chr21:1-48,129,895 | (21)x3 | 48129.9 | Pathogenic | Down syndrome | Concordant | NP |
| 4 | S-0021 | LR | 1st | NA | chr22:1 - 51,304,566 | (22)x3 | 51304.6 | Pathogenic | Trisomy 22 | NP | NP |
| 5 | S-0023 | LR | 2nd | NA | chrX:1-154,760,280 | Xp22.33q28(1_154760280)x2 | 154760.3 | Pathogenic | Klinefelter syndrome | Concordant | NP |
| 6 | S-0032 | LR | 1st | NA | chrX:1-155,270,560 | Xp22.33q28(1_154760280)x1 | 154760.3 | Pathogenic | Turner syndrome | Concordant | NP |
| 7 | S-0054 | LR | 2nd | NIPT+ | chr21:1-48,129,895 | (21)x3 | 48129.9 | Pathogenic | Down syndrome | Concordant | NP |
| 8 | S-0056 | LR | 2nd | USG+ | chrX:1-155,270,560 | Xp22.33q28(1_154760280)x1 | 154760.3 | Pathogenic | Turner syndrome | NP | NP |
| 9 | S-0058 | LR | 2nd | NIPT+ | chr21:1-48,129,895 | (21)x3 | 48129.9 | Pathogenic | Down syndrome | Concordant | NP |
| 10 | S-0062 | LR | 1st | NA | chr15:20,015,697-102,531,392 | 15q11.1q26.3(20015697_102531392)x3 | 82515.7 | Pathogenic | Partial Trisomy 15 | CBT | NP |
| 11 | S-0070 | LR | 2nd | Maternal serum+ | chr21:1-48,129,895 | (21)x3 | 48129.9 | Pathogenic | Down syndrome | Concordant | NP |
| 12 | S-0076 | LR | 2nd | USG+ | chr21:1-48,129,895 | (21)x3 | 48129.9 | Pathogenic | Down syndrome | Concordant | NP |
| 13 | S-0096 | LR | 1st | NA | chr22:16,527,284-51,304,566 | 22q11.1q13.33(16527284_51304566)x3 | 34777.3 | Pathogenic | 22q duplication syndrome | CBT | NP |
| 14 | S-0097 | LR | 1st | NA | chr15:20,015,697-102,531,392 | 15q11.1q26.3(20015697_102531392)x3 | 82515.7 | Pathogenic | Partial Trisomy 15 | CBT | NP |

| | | | | | | | | | | | |
|---|---|---|---|---|---|---|---|---|---|---|---|
| 15 | S-0098 | LR | 1st | NA | chr16:1-90,354,753 | (16)x3 | 90354.8 | Pathogenic | Trisomy 16 | Partially concordant | NP |
| | | | | | chrX:1-155,270,560 | Xp22.33q28(1_154760280)x1 | 154760.3 | Pathogenic | Turner syndrome | | |
| 16 | S-0105 | HR | 1st | USG+ | chr18:1-78,077,248 | (18)x3 | 78077.2 | Pathogenic | Edwards syndrome | Concordant | NP |
| 17 | S-0108 | HR | 2nd | USG+ | chr5:172,652,631-172,662,630 | 5q35.1(172652631_172662630)x1 | 10 | Pathogenic | Congenital Heart Defect | CBT | NP |
| 18 | S-0110 | LR | 2nd | Maternal serum+ | chr21:1-48,129,895 | (21)x3 | 48129.9 | Pathogenic | Down syndrome | NP | Concordant |
| 19 | S-0115 | LR | 1st | USG+ | chr19:7,064,492-14,814,491 | 19p13.2p13.12(7064492_14814491)x1~2 | 7750 | Pathogenic | 19p13.2 microdeletion | CBT | NP |
| 20 | S-0122 | HR | 1st | NA | chr16:1-90,354,753 | (16)x3 | 90173.6 | Pathogenic | Trisomy 16 | CBT | NP |
| 21 | S-0129 | LR | 2nd | USG+ | chr13:1-115,169,878 | (13)x3 | 115169.9 | Pathogenic | Patau Syndrome | Concordant | NP |
| 22 | S-0136 | HR | 2nd | USG+ | chr15:23,600,697-28,660,696 | 15q11.2q13.1(23600697_28660696)x3 | 5060 | Pathogenic | 15q11q13 duplication syndrome | CBT | NP |
| 23 | S-0141 | HR | 2nd | USG+ | chr18:1-78,077,248 | (18)x3 | 78077.2 | Pathogenic | Edwards syndrome | NP | Concordant |
| 24 | S-0153 | HR | 2nd | Maternal serum+ | chr21:1-48,129,895 | (21)x3 | 48129.9 | Pathogenic | Down syndrome | Concordant | NP |
| 25 | S-0177 | LR | 2nd | Maternal serum+ | chrX:1-154,760,280 | Xp22.33q28(1_154760280)x2 | 154760.3 | Pathogenic | Klinefelter syndrome | Concordant | NP |
| 26 | S-0186 | LR | 1st | NA | chr5:1-180,915,260 | (5)x3 | 180915.3 | Pathogenic | Trisomy 5 | NP | NP |
| 27 | S-0188 | LR | 2nd | USG+ | chr21:1-48,129,895 | (21)x3 | 48129.9 | Pathogenic | Down syndrome | Concordant | NP |
| 28 | S-0200 | LR | 1st | NA | chr21:1-48,129,895 | (21)x3 | 38564.9 | Pathogenic | Down syndrome | Concordant | NP |
| 29 | S-0207 | HR | 2nd | USG+ | chr21:1-48,129,895 | (21)x3 | 48129.9 | Pathogenic | Down syndrome | Concordant | NP |

| | | | | | | | | | | | |
|---|---|---|---|---|---|---|---|---|---|---|---|
| 30 | S-0222 | HR | 1st | NA | chr16:1-90,354,753 | (16)x3 | 90354.8 | Pathogenic | Trisomy 16 | CBT | NP |
| 31 | S-0226 | LR | 2nd | USG+ | chr21:1-48,129,895 | (21)x3 | 48129.9 | Pathogenic | Down syndrome | NP | Concordant |
| 32 | S-0240 | HR | 2nd | Maternal serum+ | chr22:18,646,034-21,713,533 | 22q11.21(18646034_21713533)x1 | 3067.5 | Pathogenic | 22q11.21 deletion syndrome | CBT | NP |
| 33 | S-0248 | LR | 1st | NA | chrX:1-155,270,560 | Xp22.33q28(1_154760280)x1 | 154760.3 | Pathogenic | Turner syndrome | NP | NP |
| 34 | S-0276 | LR | 2nd | NIPT+ | chr21:1-48,129,895 | (21)x3 | 48129.9 | Pathogenic | Down syndrome | Concordant | NP |
| 35 | S-0285 | LR | 2nd | NA | chrX:1-155,270,560 | Xp22.33q28(1_154760280)x3 | 154760.3 | Pathogenic | Triple X syndrome | Concordant | NP |
| 36 | S-0291 | HR | 1st | USG+ | chr18:1-78,077,248 | (18)x3 | 78077.2 | Pathogenic | Edwards syndrome | Concordant | NP |
| 37 | S-0297 | HR | 1st | USG+ | chr21:1-48,129,895 | (21)x3 | 48129.9 | Pathogenic | Down syndrome | Concordant | NP |
| 38 | S-0298 | HR | 2nd | USG+ | chr22:20,706,034-21,641,033 | 22q11.21(20706034_21641033)x1 | 935 | Likely Pathogenic | 22q11.21 deletion | CBT | NP |
| 39 | S-0309 | LR | 2nd | USG+ | chr21:1-48,129,895 | (21)x3 | 48129.9 | Pathogenic | Down syndrome | Concordant | NP |
| 40 | S-0310 | LR | 2nd | USG+ | chr22:23,777,284-25,027,283 | 22q11.23(23777284_25027283)x3 | 1250 | VUS | Distal 22q11.2 microduplication syndrome | CBT | NP |
| 41 | S-0314 | HR | 1st | NA | All chromosomes | 69, XXY | - | Pathogenic | Triploidy | Concordant | NP |
| 42 | S-0317 | LR | 2nd | NIPT+ | chr21:1-48,129,895 | (21)x3 | 48129.9 | Pathogenic | Down syndrome | Concordant | NP |
| 43 | S-0335 | LR | 1st | NA | chr16:1-1,802,376 | 16p13.3(1_1802376)x1 | 1802.4 | Pathogenic | 16p13.3 deletion syndrome | CBT | NP |
| | | | | | chr11:113,253,259-135,006,516 | 11q23.2q25(113253259_135006516)x3 | 21753.3 | Likely Pathogenic | 11q23.2q25 duplication syndrome | | |

| | | | | | | | | | | | |
|---|---|---|---|---|---|---|---|---|---|---|---|
| 44 | S-0345 | LR | 2nd | USG+ | chr21:1-48,129,895 | (21)x3 | 48129.9 | Pathogenic | Down syndrome | Concordant | NP |
| 45 | S-0346 | LR | 2nd | USG+ | chr21:1-48,129,895 | (21)x3 | 48129.9 | Pathogenic | Down syndrome | Concordant | NP |
| 46 | S-0356 | LR | 2nd | USG+ | chr21:1-48,129,895 | (21)x3 | 48129.9 | Pathogenic | Down syndrome | Partially concordant | NP |
| | | | | | chr1:146,468,726-148,025,293 | 1q21.1q21.2(145750311_148250310)x3 | 1556.6 | Pathogenic | 1q21.1q21.2 microduplication syndrome | | |
| 47 | S-0364 | HR | 1st | USG+ | chr18:1-78,077,248 | (18)x3 | 78077.2 | Pathogenic | Edwards Syndrome | Concordant | NP |
| 48 | S-0376 | LR | 1st | NA | chr13:1-115,169,878 | (13)x3 | 115169.9 | Pathogenic | Patau syndrome | Concordant | NP |
| 49 | S-0381 | LR | 2nd | USG+ | chr22:18,777,284-21,527,283 | 22q11.21(18777284_21527283)x1 | 2750 | Pathogenic | 22q11.21 deletion syndrome | CBT | NP |
| 50 | S-0395 | LR | 2nd | NA | chr22:18,889,102-21,510,720 | 22q11.21(18889102_21510720)x3 | 2621 | Pathogenic | 22q11.21 duplication syndrome | CBT | NP |
| 51 | S-0415 | LR | 1st | NA | All chromosomes | 23,X0 | - | Pathogenic | Haploidy | Concordant | NP |
| 52 | S-0421 | HR | 2nd | USG+ | chrX:31,832,781-31,907,780 | Xp21.1(31832781_31907780)x0 | 75 | Pathogenic | Duchenne Muscular Dystrophy (DMD) | CBT | NP |
| 53 | S-0428 | LR | 2nd | USG+ | chrX:1-155,270,560 | Xp22.33q28(1_154760280)x1 | 154760.3 | Pathogenic | Turner syndrome | Concordant | Concordant |
| 54 | S-0435 | LR | 2nd | USG+ | chr21:1-48,129,895 | (21)x3 | 48129.9 | Pathogenic | Down syndrome | Concordant | NP |
| 55 | S-0440 | LR | 2nd | NIPT+ | chr18:1-78,077,248 | (18)x3 | 78077.248 | Pathogenic | Edwards Syndrome | Concordant | NP |

| | | | | | | | | | | | |
|---|---|---|---|---|---|---|---|---|---|---|---|
| 56 | S-0455 | HR | 2nd | USG+ | chr2:166,853,437-166,868,436 | 2q24.3(166853437_166868436)x1 | 15 | Likely Pathogenic | Dravet syndrome/Febrile seizures, familial, 3A/ Generalized epilepsy with febrile seizures plus, type 2 | CBT | NP |
| 57 | S-0468 | HR | 2nd | USG+ | chr2:130,830,001-131,194,000 | 2q21.1(130830001_131194000)x1 | 364 | Pathogenic | 2q21.1 microdeletion | CBT | NP |
| 58 | S-0499 | HR | 2nd | USG+ | chr7:143,773,082-159,138,663 | 7q35q36.3(143773082_159138663)x3 | 15365.6 | Pathogenic | 7q35q36.3 duplication syndrome | CBT | Concordant |
| | | | | | chr18:72,408,625-78,077,248 | 18q22.3q23(72408625_78077248)x1 | 5668.8 | Likely Pathogenic | 18q22.3q23 deletion syndrome | | |
| 59 | S-0502 | HR | 2nd | USG+ | chr21:1-48,129,895 | (21)x3 | 48129.9 | Pathogenic | Down syndrome | Concordant | NP |
| 60 | S-0507 | LR | 1st | USG+ | chr21:1-48,129,895 | (21)x3 | 48129.9 | Pathogenic | Down syndrome | Concordant | NP |
| 61 | S-0516 | LR | 2nd | USG+ | chr17:34,847,606-36,097,605 | 17q12(34847606_36097605)x1 | 1250 | Pathogenic | 17q12 deletion syndrome | CBT | NP |
| 62 | S-0524 | LR | 2nd | USG+ | chr21:1-48,129,895 | (21)x3 | 48129.9 | Pathogenic | Down syndrome | Concordant | NP |
| 63 | S-0527 | LR | 2nd | USG+ | chr1:144,750,311-149,500,310 | 1q21.1q21.2(144750311_149500310)x3 | 4750 | Pathogenic | 1q21.1q21.2 duplication syndrome | CBT | NP |
| 64 | S-0544 | LR | 2nd | USG+ | chr22:23,027,284-23,777,283 | 22q11.22q11.23(23027284_23777283)x1 | 750 | Pathogenic | 22q11.22q11.23 deletion syndrome | CBT | NP |
| 65 | S-0545 | LR | 2nd | USG+ | chr16:28,302,377-30,302,376 | 16p11.2(28302377_30302376)x1 | 2000 | Pathogenic | 16p11.2 deletion syndrome | CBT | NP |
| 66 | S-0587 | LR | 1st | NA | chr10:1-135,534,747 | (10)x3 | 135534.7 | Pathogenic | Trisomy 10 | CBT | NP |

| | | | | | | | | | | | |
|---|---|---|---|---|---|---|---|---|---|---|---|
| 67 | S-0608 | LR | 2nd | USG+ | chrY:1-59,373,566 | Yp11.32q12(1_28811783)x2 | 59373.6 | Pathogenic | Jacobs syndrome | Concordant | NP |
| 68 | S-0611 | LR | 2nd | USG+ | chrX:1-154,760,280 | Xp22.33q28(1_154760280)x2 | 154760.3 | Pathogenic | Klinefelter syndrome | Concordant | NP |
| 69 | S-0634 | HR | 2nd | USG+ | chr1:146,467,811-147,907,810 | 1q21.1q21.2(146467811_147907810)x3 | 1440 | Pathogenic | 1q21.1q21.2 microduplication syndrome | CBT | NP |
| 70 | S-0648 | LR | 1st | NA | chr21:1-48,129,895 | (21)x3 | 48129.9 | Pathogenic | Down syndrome | Concordant | NP |
| 71 | S-0687 | LR | 2nd | USG+ | chr18:1-78,077,248 | (18)x3 | 78077.2 | Pathogenic | Edwards Syndrome | Concordant | NP |
| 72 | S-0692 | LR | 2nd | Maternal serum+ | chr21:1-48,129,895 | (21)x3 | 48129.9 | Pathogenic | Down syndrome | Concordant | NP |
| 73 | S-0696 | HR | 2nd | NA | chr1:145,828,001-147,900,000 | 1q21.1q21.2(145828001_147900000)x3 | 2072 | Likely Pathogenic | 1q21.1q21.2 microduplication syndrome | CBT | NP |
| 74 | S-0699 | LR | 1st | NA | chrX:1-155,270,560 | Xp22.33q28(1_154760280)x1 | 154760.3 | Pathogenic | Turner syndrome | Concordant | NP |
| 75 | S-0700 | LR | 2nd | NIPT+ | chr21:1-48,129,895 | (21)x3 | 48129.9 | Pathogenic | Down syndrome | Concordant | NP |
| 76 | S-0712 | LR | 2nd | USG+ | chr21:1-48,129,895 | (21)x3 | 48129.9 | Pathogenic | Down syndrome | Concordant | NP |
| 77 | S-0724 | LR | 2nd | USG+ | chr21:1-48,129,895 | (21)x3 | 48129.9 | Pathogenic | Down syndrome | Concordant | NP |
| 78 | S-0729 | HR | 1st | USG+ | chr13:1-115,169,878 | (13)x3 | 115169.9 | Pathogenic | Patau syndrome | Concordant | NP |
| 79 | S-0730 | LR | 2nd | Maternal serum+ | chr21:1-48,129,895 | (21)x3 | 48129.9 | Pathogenic | Down syndrome | Concordant | NP |
| 80 | S-0744 | HR | 1st | NA | chr21:1-48,103,697 | 21p13q22.3(1_48103697)x1 | 48103.7 | Pathogenic | Monosomy 21 | Concordant | NP |
| 81 | S-0758 | HR | 1st | NA | chr2:1-243,199,373 | (2)x3 | 243199.4 | Pathogenic | Trisomy 2 | CBT | NP |
| 82 | S-0779 | LR | 2nd | USG+ | chr16:15,052,377-16,302,376 | 16p13.11(15052377_16302376)x1 | 1250 | Pathogenic | 16p13.11 microdeletion syndrome | CBT | NP |

| | | | | | | | | | | | |
|---|---|---|---|---|---|---|---|---|---|---|---|
| 83 | S-0780 | LR | 2nd | USG+ | chr9:1-10,856,715 | 9p24.3p23(1_10856715)x3 | 10856.7 | VUS | 9p24.3p23 duplication syndrome | CBT | NP |
| | | | | | chr14:19,049,771-47,549,770 | 14q11.1q21.3(19049771_47549770)x3 | 28500 | Pathogenic | 14q11.2q21.3 duplication syndrome | | |
| 84 | S-0781 | LR | 2nd | USG+ | chr18:1-78,077,248 | (18)x3 | 78077.2 | Pathogenic | Edwards Syndrome | Concordant | NP |
| 85 | S-0786 | LR | 2nd | USG+ | chr21:1-48,129,895 | (21)x3 | 48129.9 | Pathogenic | Down syndrome | Concordant | NP |
| 86 | S-0791 | LR | 2nd | USG+ | chr21:1-48,129,895 | (21)x3 | 48129.9 | Pathogenic | Down syndrome | Concordant | NP |
| 87 | S-0801 | LR | 2nd | USG+ | chr21:1-48,129,895 | (21)x3 | 48129.9 | Pathogenic | Down syndrome | Concordant | NP |
| 88 | S-0806 | HR | 2nd | NIPT+ | chrY:8,403,034-59,353,033 | Yp11.2q12(8403034_59353033)x0 | 50950 | Pathogenic | Yp11.2q12 deletion syndrome | NP | NP |
| 89 | S-0816 | LR | 2nd | Maternal serum+ | chr21:1-48,129,895 | (21)x3 | 48129.9 | Pathogenic | Down syndrome | Concordant | NP |
| 90 | S-0834 | LR | 2nd | NA | All Chromosomes | 69, XXY | - | Pathogenic | Triploidy | Concordant | NP |
| 91 | S-0836 | LR | 2nd | USG+ | chr15:22,765,697-23,265,696 | 15q11.2(22765697_23265696)x1 | 500 | VUS | 15q11.2 (BP1-BP2) microdeletion syndrome | CBT | NP |
| 92 | S-0844 | LR | 2nd | USG+ | chr21:1-48,129,895 | (21)x3 | 48129.9 | Pathogenic | Down syndrome | Concordant | NP |
| 93 | S-0860 | HR | 2nd | USG+ | chr21:1-48,129,895 | (21)x3 | 48129.9 | Pathogenic | Down syndrome | Concordant | NP |
| 94 | S-0875 | HR | 1st | NA | chr21:1-48,129,895 | (21)x3 | 48129.9 | Pathogenic | Down syndrome | Concordant | NP |
| 95 | S-0881 | LR | 1st | NA | chrX:1-155,270,560 | Xp22.33q28(1_154760280)x1 | 154760.3 | Pathogenic | Turner syndrome | Concordant | NP |
| 96 | S-0904 | LR | 1st | USG+ | chr22:16,527,284-51,304,566 | 22q11.1q13.33(16527284_51304566)x3 | 34777.3 | Pathogenic | 22q duplication syndrome | CBT | NP |

| | | | | | | | | | | | |
|---|---|---|---|---|---|---|---|---|---|---|---|
| 97 | S-0906 | LR | 2nd | USG+ | chr21:1-48,129,895 | (21)x2~3 | 48129.9 | Pathogenic | Down syndrome | BDL | NP |
| 98 | S-0914 | LR | 1st | USG+ | chrX:1-155,270,560 | Xp22.33q28(1_154760280)x1 | 154760.3 | Pathogenic | Turner syndrome | Concordant | NP |
| 99 | S-0916 | LR | 2nd | USG+ | chr17:34,847,606-36,347,605 | 17q12(34847606_36347605)x1 | 1500 | Pathogenic | 17q12 deletion syndrome | CBT | NP |
| 100 | S-0927 | HR | 1st | USG+ | chr21:1-48,129,895 | (21)x3 | 48129.9 | Pathogenic | Down syndrome | Concordant | NP |
| 101 | S-0935 | LR | 2nd | USG+ | chr22:22,980,871-23,630,834 | 22q11.22q11.23(23027284_23527283)x1 | 500 | VUS | 22q11.22q11.23 deletion syndrome | CBT | NP |
| 102 | S-0937 | LR | 2nd | USG+ | chr21:1-48,129,895 | (21)x3 | 48129.9 | Pathogenic | Down syndrome | Concordant | NP |
| 103 | S-0939 | LR | 2nd | NA | chrX:1-155,270,560 | Xp22.33q28(1_154760280)x1 | 154760.3 | Pathogenic | Turner syndrome | Concordant | NP |
| 104 | S-0948 | LR | 2nd | NIPT+ | chr21:1-48,129,895 | (21)x3 | 48129.9 | Pathogenic | Down syndrome | Concordant | NP |
| 105 | S-0952 | LR | 2nd | NIPT+ | chrX:1-155,270,560 | Xp22.33q28(1_154760280)x3 | 154760.3 | Pathogenic | Triple X syndrome | Concordant | NP |
| 106 | S-0967 | LR | 2nd | USG+ | chr21:1-48,129,895 | (21)x3 | 48129.9 | Pathogenic | Down syndrome | Concordant | NP |
| 107 | S-0975 | LR | 2nd | USG+ | chr21:1-48,129,895 | (21)x3 | 48129.9 | Pathogenic | Down syndrome | Concordant | NP |
| 108 | S-0994 | LR | 2nd | Maternal serum+ | chr18:1-78,077,248 | (18)x3 | 78077.2 | Pathogenic | Edwards Syndrome | Concordant | NP |
| 109 | S-1005 | HR | 1st | NA | chr22:16,366,034-51,304,566 | 22q11.1q13.33(16366034_51304566)x3 | 34777.3 | Pathogenic | 22q duplication syndrome | CBT | NP |
| 110 | S-1011 | LR | 2nd | USG+ | chr22:18844591-21518340 | 22q11.21(18844591_21518340)x3 | 2673.8 | Pathogenic | 22q11.21 duplication syndrome | CBT | NP |
| 111 | S-1027 | LR | 1st | NA | chr10:1-135,534,747 | (10)x3 | 135534.7 | Pathogenic | Trisomy 10 | CBT | NP |
| 112 | S-1032 | LR | 2nd | Maternal serum+ | chr18:1-78,077,248 | (18)x3 | 78077.2 | Pathogenic | Edwards Syndrome | Concordant | NP |

| | | | | | | | | | | | |
|---|---|---|---|---|---|---|---|---|---|---|---|
| 113 | S-1054 | LR | 2nd | NIPT+ | chrX:1-155,270,560 | Xp22.33q28(1_154760280)x1 | 154760.3 | Pathogenic | Turner syndrome | Concordant | NP |
| 114 | S-1057 | LR | 1st | NA | chrY:1-59,373,566 | (Y)x1~2 | 59373.6 | Pathogenic | Jacobs Syndrome | BDL | NP |
| 115 | S-1059 | LR | 1st | NA | chr18:1-78,077,248 | (18)x3 | 78077.2 | Pathogenic | Edwards Syndrome | Concordant | NP |
| 116 | S-1065 | LR | 2nd | USG+ | chr21:1-48,129,895 | (21)x3 | 48129.9 | Pathogenic | Down syndrome | Concordant | NP |
| 117 | S-1079 | LR | 2nd | USG+ | chr21:1-48,129,895 | (21)x3 | 48129.9 | Pathogenic | Down syndrome | Concordant | NP |
| 118 | S-1091 | LR | 1st | NA | chr17:1-81195210 | (17)x3 | 81171.8 | Pathogenic | Trisomy 17 | CBT | NP |
| 119 | S-1094 | LR | 1st | NA | chr4:1-191,154,276 | (4)x3 | 191154.3 | Pathogenic | Trisomy 4 | CBT | NP |
| 120 | S-1101 | LR | 2nd | USG+ | chrX:1-154,760,280 | Xp22.33q28(1_155270560)x2 | 155270.5 | Pathogenic | Klinefelter syndrome | Concordant | NP |
| 121 | S-1103 | LR | 3rd | NIPT+ | chr21:1-48,129,895 | (21)x3 | 48129.9 | Pathogenic | Down syndrome | Concordant | NP |
| 122 | S-1126 | LR | 2nd | USG+ | chr18:1-78,077,248 | (18)x3 | 78077.2 | Pathogenic | Edwards Syndrome | Concordant | NP |
| 123 | S-1135 | LR | 2nd | NA | chr16:5,052,377-35,302,376 | 16p13.3p11.1(5052377_35302376)x2~3 | 30250 | Pathogenic | 16p13.3 p11.1 duplication syndrome | NP | NP |
| | | | | | chr19:1-59,128,983 | (19)x2~3 | 59129 | Likely Pathogenic | Trisomy 19 | | |
| | | | | | chrX:1-155,270,560 | Xp22.33q28(1_154760280)x3 | 154760.3 | Pathogenic | Triple X syndrome | | |
| 124 | S-1147 | LR | 2nd | USG+ | chr7:148270816-159138663 | 7q36.1-q36.3(148270816_159138663)x1 | 10867.8 | Pathogenic | 7q36.1 q36.3 deletion syndrome | CBT | NP |
| | | | | | chr6:171165-28156478 | 6p25.3-p22.1(171165_28156478)x3 | 27985.3 | Pathogenic | 6p25.3 p22.1 duplication syndrome | | |
| 125 | S-1151 | LR | 2nd | USG+ | chr17:1-2,036,784 | 17p13.3(1_2036784)x1 | 2036.7 | Pathogenic | 17p13.3 microdeletion syndrome | CBT | NP |

| | | | | | | | | | | | |
|---|---|---|---|---|---|---|---|---|---|---|---|
| 126 | S-1161 | HR | 2nd | Maternal serum+ | chr21:1-48,129,895 | (21)x3 | 48129.9 | Pathogenic | Down syndrome | Concordant | NP |
| 127 | S-1165 | LR | 1st | USG+ | chr4:90,953-15,279,936 | 4p16.3p15.32(90953_15279936)x1 | 15188.9 | Pathogenic | Wolf-Hirschhorn syndrome | Concordant | NP |
| | | | | | chr8:181,905-17,417,308 | 8p23.3p22(181905_17417308)x3 | 17235.4 | Pathogenic | 8p23.3p22 duplication syndrome | | NP |
| 128 | S-1186 | LR | 2nd | USG+ | chr21:1-48,129,895 | (21)x3 | 48129.9 | Pathogenic | Down syndrome | Concordant | NP |
| 129 | S-1195 | LR | 2nd | USG+ | chr21:1-48,129,895 | (21)x3 | 48129.9 | Pathogenic | Down syndrome | Concordant | NP |
| 130 | S-1199 | LR | 2nd | Maternal serum+ | chrX:1-155,270,560 | Xp22.33q28(1_155270560)x3 | 155270.5 | Pathogenic | Triple X syndrome | Concordant | NP |
| 131 | S-1207 | LR | 1st | NA | chr21:1-48,129,895 | (21)x3 | 48129.9 | Pathogenic | Down syndrome | Concordant | NP |
| 132 | S-1212 | LR | 1st | NA | chrX:1-155,270,560 | Xp22.33q28(1_154760280)x1 | 154760.3 | Pathogenic | Turner syndrome | Concordant | NP |
| 133 | S-1226 | LR | 2nd | Maternal serum+ | chr4:178074941-190980220 | 4q34.3q35.2(178074941_190980220)x1 | 12905.2 | Pathogenic | 4q34.3q35.2 deletion syndrome | CBT | NP |
| | | | | | chr11:117357391-134967720 | 11q23.3q25(117357391_134967720)x3 | 17610.3 | Pathogenic | 11q23.3q25 duplication syndrome | | |
| 134 | S-1238 | LR | 2nd | NIPT+ | chr18:1-78,077,248 | (18)x3 | 78077.2 | Pathogenic | Edwards Syndrome | Concordant | NP |
| 135 | S-1250 | LR | 2nd | USG+ | chr21:1-48,129,895 | (21)x3 | 48129.9 | Pathogenic | Down syndrome | Concordant | NP |

| | | | | | | | | | | | |
|---|---|---|---|---|---|---|---|---|---|---|---|
| 136 | S-1253 | LR | 2nd | USG+ | chr3:136906849-141004800 | 3q22.3q23(136906849_141004800)x1 | 4097.9 | Pathogenic | Blepharophimosis-Ptosis-Epicanthus Inversus Syndrome (BPES) | CBT | NP |
| 137 | S-1265 | LR | 1st | NA | chr16:1-90354753 | (16)x3 | 90354.7 | Pathogenic | Trisomy 16 | CBT | NP |
| 138 | S-1285 | LR | 1st | NA | chrY:1-59,373,566 | (Y)x0~1 | 59373.5 | Pathogenic | Mosaic loss of the Y Chromosome (mLOY) | BDL | NP |
| 139 | S-1317 | HR | 1st | NA | chr21:1-48,129,895 | (21)x1 | 48129.8 | Pathogenic | Monosomy 21 | Concordant | NP |
| 140 | S-1319 | LR | 1st | NA | chrX:1-155,270,560 | Xp22.33q28(1_154760280)x1 | 154760.3 | Pathogenic | Turner syndrome | Concordant | NP |
| 141 | S-1322 | LR | 2nd | USG+ | chr21:1-48,129,895 | (21)x3 | 48129.9 | Pathogenic | Down syndrome | Concordant | NP |
| 142 | S-1325 | LR | 2nd | USG+ | chr21:1-48,129,895 | (21)x3 | 48129.9 | Pathogenic | Down syndrome | Concordant | NP |
| 143 | S-1338 | LR | 1st | NA | chrX:1-155,270,560 | Xp22.33q28(1_154760280)x1 | 154760.3 | Pathogenic | Turner syndrome | Concordant | NP |
| 144 | S-1341 | LR | 1st | NA | chrX:1-155,270,560 | Xp22.33q28(1_154760280)x1 | 154760.3 | Pathogenic | Turner syndrome | Concordant | NP |
| | | | | | chr13:1-115,169,878 | (13)x2~3 | 115169.9 | Pathogenic | Patau syndrome | | |
| 145 | S-1349 | LR | 1st | NA | chrX:1-155,270,560 | Xp22.33q28(1_155270560)x2~3 | 155270.5 | Pathogenic | Triple X syndrome | BDL | NP |
| 146 | S-1350 | LR | 2nd | NIPT+ | chr9:1-47,106,715 | 9p24.3p11.2(1_47106715)x2~3 | 47106.7 | Pathogenic | 9p24.3p11.2 duplication syndrome | CBT | NP |
| 147 | S-1351 | LR | 2nd | USG+ | chr21:1-48,129,895 | (21)x3 | 48129.9 | Pathogenic | Down syndrome | Concordant | NP |

| | | | | | | | | | | | |
|---|---|---|---|---|---|---|---|---|---|---|---|
| 148 | S-1360 | LR | 2nd | USG+ | chr21:1-48,129,895 | (21)x3 | 48129.9 | Pathogenic | Down syndrome | Concordant | NP |
| 149 | S-1370 | LR | 2nd | USG+ | chr15:22636440-23449468 | 15q11.2(22636440_23449468)x1 | 813 | VUS | 15q11.2 (BP1-BP2) microdeletion syndrome | CBT | NP |
| 150 | S-1371 | LR | 2nd | USG+ | chr21:1-48,129,895 | (21)x3 | 48129.9 | Pathogenic | Down syndrome | Concordant | NP |
| 151 | S-1372 | LR | 2nd | Maternal serum+ | chr18:1-78,077,248 | (18)x3 | 78077.2 | Pathogenic | Edwards Syndrome | Concordant | NP |
| 152 | S-1387 | LR | 2nd | USG+ | chr21:1-48,129,895 | (21)x3 | 48129.9 | Pathogenic | Down syndrome | Concordant | NP |
| 153 | S-1389 | LR | 2nd | USG+ | chr18:1-78,077,248 | (18)x3 | 78077.2 | Pathogenic | Edwards Syndrome | Concordant | NP |
| 154 | S-1415 | LR | 1st | NA | All Chromosomes | 69, XXY | - | Pathogenic | Triploidy | Concordant | NP |
| 155 | S-1420 | LR | 2nd | USG+ | chr21:1-48,129,895 | (21)x3 | 48129.9 | Pathogenic | Down syndrome | Concordant | NP |
| 156 | S-1438 | LR | 2nd | USG+ | chr22:18630406-21712472 | 22q11.21(18630406_21712472)x1 | 3082 | Pathogenic | 22q11.21 Deletion Syndrome | CBT | NP |
| 157 | S-1443 | LR | 2nd | Maternal serum+ | chr21:1-48,129,895 | (21)x3 | 48129.9 | Pathogenic | Down syndrome | Concordant | NP |
| 158 | S-1447 | LR | 2nd | USG+ | chr17:34752628-36322648 | 17q12q12(34752628_36322648)x1 | 1570 | Pathogenic | 17q12 deletion syndrome | CBT | NP |
| 159 | S-1479 | LR | 1st | USG+ | chr21:1-48,129,895 | (21)x3 | 48129.9 | Pathogenic | Down syndrome | Concordant | NP |
| 160 | S-1483 | LR | 2nd | USG+ | chr11:45871953-47241264 | 11p11.2(45871953_47241264)x1 | 1369.3 | Pathogenic | Potocki-Shaffer syndrome (PSS) | CBT | Concordant |
| 161 | S-1497 | LR | 2nd | NIPT+ | chr21:1-48,129,895 | (21)x3 | 48129.9 | Pathogenic | Down syndrome | Concordant | NP |

| | | | | | | | | | | | |
|---|---|---|---|---|---|---|---|---|---|---|---|
| 162 | S-1498 | LR | 2nd | USG+ | chrX:128374-55542718 | Xp22.33p11.21(128374_55542718)x1 | 55414.345 | Pathogenic | Xp22.33 p11.21 deletion | CBT | NP |
| | | | | | chrX:123109708-155270560 | Xq25q28(123109708_155270560)x3 | 32160.853 | Pathogenic | Xq25 q28 duplication | | |
| 163 | S-1503 | LR | 2nd | USG+ | chr21:1-48,129,895 | (21)x3 | 48129.9 | Pathogenic | Down syndrome | Concordant | NP |
| 164 | S-1504 | HR | 2nd | NA | chr4:179,233,389-191,154,276 | 4q34.3q35.2(179233389_191154276)x1 | 11920.8 | Pathogenic | 4q34.3 q35.2 Deletion | CBT | NP |
| | | | | | chr8:125,762-31,355,761 | 8p23.3p12(125762_31355761)x3 | 31230 | Pathogenic | 8p23.3 p12 Duplication | | |
| 165 | S-1505 | LR | 2nd | USG+ | chr9:130,760,129-141,114,824 | 9q34.11q34.3(130760129_141114824)x3 | 10354.6 | Pathogenic | 9q34.11q34.3 duplication syndrome | CBT | NP |
| 166 | S-1506 | LR | 2nd | USG+ | chr15:22592740-28939014 | 15q11.2q13.1(22592740_28939014)x3 | 6346.2 | Pathogenic | 15q11q13 micro duplication syndrome | CBT | NP |
| 167 | S-1507 | LR | 2nd | USG+ | chr21:1-48,129,895 | (21)x3 | 48129.8 | Pathogenic | Down syndrome | Concordant | NP |

| | | | | | | | | | | | |
|---|---|---|---|---|---|---|---|---|---|---|---|
| 168 | S-1508 | LR | 1st | NA | chr10:1-135,534,747 | (10)x2~3 | 135534.7 | Pathogenic | Trisomy 10 | CBT | NP |

**Abbreviations:**
LR - Low resolution; HR - High resolution; USG - Ultrasonography; NIPT - Non-invasive prenatal testing; NA - Not available; VUS - Variant of uncertain significance; NP - Not Performed; BDL - Below detection limit; CBT - Cannot be tested

**Supplementary Table S2: Prevalence of USG soft markers**

| USG soft markers | Total number of cases with the soft marker | No. of cases with positive finding |
|---|---|---|
| NT/NFT | 187 | 35 |
| AHNB | 210 | 37 |
| EIF | 143 | 20 |
| EB | 32 | 7 |
| Ventriculomegaly | 49 | 3 |
| ARSA | 54 | 5 |
| VSD | 37 | 9 |

**Abbreviations:**
NT/NFT -Nuchal translucency/Nuchal fold thickness; AHNB - Absent or hypoplastic nasal bone; EIF - Echogenic intracardiac focus; EB - Echogenic bowel; ARSA - Aberrant right subclavian artery; VSD - Ventricular septal defect; USG - Ultrasonography

**Supplementary Table S3: USG soft markers observed in commonly reported syndromes in this study (excluding POC samples)**

| | NT/NFT | AHNB | EIF | Ventriculomegaly | VSD | ARSA | EB | Other common USG findings |
|---|---|---|---|---|---|---|---|---|
| Down syndrome (54) | 17 | 22 | 15 | 2 | 6 | 4 | 4 | Tricuspid regurgitation, ductus venosus agenesis, AVSD and increased prenasal thickness |
| Edwards syndrome (9) | 1 | 4 | - | - | - | - | 2 | Congenital heart disease, agenesis of corpus callosum, omphalocele, ganglion eminence cyst |
| Klinefelter (4) | - | - | 1 | - | - | - | - | - |
| Triple X syndrome (3) | - | 1 | - | - | - | - | - | - |
| Patau syndrome (1) | 1 | - | - | - | - | - | - | Hydrops fetalis,dysmorphic facial appearance |
| Turner syndrome (1) | 1 | - | - | - | - | - | - | Cystic hygroma |
| Jacobs syndrome (1) | - | - | 1 | 1 | - | - | - | _ |
| Microdeletions / Microduplications (46) | 7 | 7 | 3 | - | 2 | 1 | 1 | Cardiac abnormality- right aortic arch, persistent left SVC, TOF; dysmorphic facial features, echogenic kidneys. |

**Abbreviations:**
NT/NFT -Nuchal translucency/Nuchal fold thickness; AHNB - Absent or hypoplastic nasal bone; EIF - Echogenic intracardiac focus; EB - Echogenic bowel; ARSA - Aberrant right subclavian artery; VSD - Ventricular septal defect; AVSD - Atrio-ventricular septal defect; SVC - Superior vena cava; TOF - Tetralogy of Fallot; USG - Ultrasonography

**Supplementary Table S4: Summary of cases subjected to Whole Exome Sequencing (WES), including clinical outcomes and cases where a confirmatory diagnosis was established based on identified single nucleotide variants (SNVs).**

| S No. | Sample ID | Gene/ Genomic region | Varaint | Zygosity | Classification | MOI (Mode of Inheritance) |
|---|---|---|---|---|---|---|
| 1 | S-0001 | *PCLO* | chr7:82764767G>A<br>c.2099C>T<br>p.Pro700Leu | Heterozygous | Variant of Uncertain Significance (VUS) | Recessive |
| | | *PCLO* | chr7:82763587G>T<br>c.3279C>A<br>p.Asn1093Lys | Heterozygous | Variant of Uncertain Significance (VUS) | Recessive |
| 2 | S-0010 | *PUF60* | g.144906540_144906543delinsGAA<br>c.51_54delinsTTC<br>p.Glu19SerfsTer41 | Heterozygous | Likely Pathogenic | Dominant |
| 3 | S-0013 | Negative for disease-causing or likely disease-causing variants | | | | |
| 4 | S-0015 | *GALC* | chr14:88434707G>A<br>c.880C>T<br>p.Gln294Ter | Homozygous | Likely Pathogenic | Recessive |
| 5 | S-0025 | *BCKDHB* | chr6:80837335A>C<br>c.268A>C<br>p.Thr90Pro | Heterozygous | Variant of Uncertain Significance (VUS) | Recessive |
| 6 | S-0026 | Negative for disease-causing or likely disease-causing variants | | | | |
| 7 | S-0077 | Negative for disease-causing or likely disease-causing variants | | | | |
| 8 | S-0110 | chr21pter_qter | chr21pter_qter[3]<br>[Trisomy 21] | Inconclusive | Pathogenic | NA |
| 9 | S-0111 | Negative for disease-causing or likely disease-causing variants | | | | |
| 10 | S-0120 | Negative for disease-causing or likely disease-causing variants | | | | |
| 11 | S-0121 | Negative for disease-causing or likely disease-causing variants | | | | |
| 12 | S-0135 | Negative for disease-causing or likely disease-causing variants | | | | |
| 13 | S-0139 | Negative for disease-causing or likely disease-causing variants | | | | |
| 14 | S-0141 | chr18pter_qter | chr18pter_qter[3]<br>[Trisomy 18] | Inconclusive | Pathogenic | NA |

| | | | | | | |
|---|---|---|---|---|---|---|
| 15 | S-0144 | Negative for disease-causing or likely disease-causing variants | | | | |
| 16 | S-0163 | chr15q11.2 | chr15:(?_22833519)<br>_(23086416_?)del<br>(chr15q11.2 partial deletion) | Heterozygous | Pathogenic | Dominant |
| 17 | S-0166 | *HBA2* | chr16:(?_222911)_(223600_?)del<br>c.(?_-1)_(*1_?)del<br>[Whole gene deletion (exon 1-3)] | Homozygous | Pathogenic | Recessive |
| 18 | S-0197 | *FREM1* | chr9:14807972G>C<br>c.3054C>G<br>p.Tyr1018Ter | Heterozygous | Likely Pathogenic | Dominant/Recessive |
| 19 | S-0219 | *ADA* | chr20:43254264G>A<br>c.424C>T<br>p.Arg142Ter | Heterozygous | Pathogenic | Recessive |
| | | *PALB2* | chr16:23619279G>A<br>c.3256C>T<br>p.Arg1086Ter | Heterozygous | Pathogenic | Dominant |
| 20 | S-0226 | chr21pter_qter | chr21pter_qter[3]<br>[Trisomy 21] | Inconclusive | Pathogenic | NA |
| 21 | S-0227 | Negative for disease-causing or likely disease-causing variants | | | | |
| 22 | S-0398 | *CC2D2A* | chr4:15538701T>C<br>c.1764+2T>C | Homozygous | Likely Pathogenic | Recessive |
| 23 | S-0412 | Negative for disease-causing or likely disease-causing variants | | | | |
| 24 | S-0419 | Negative for disease-causing or likely disease-causing variants | | | | |
| 25 | S-0428 | chrXpter_qter | chrXpter_qter[1]<br>[Deletion of chromos<br>ome X] | Heterozygous | Pathogenic | NA |
| 26 | S-0454 | *ANTXR2* | chr4:80905989dupG<br>c.1073dupC<br>p.Ala359CysfsTer13 | Homozygous | Pathogenic | Recessive |
| 27 | S-0509 | Negative for disease-causing or likely disease-causing variants | | | | |
| 28 | S-0511 | Negative for disease-causing or likely disease-causing variants | | | | |
| 29 | S-0513 | Negative for disease-causing or likely disease-causing variants | | | | |

| | | | | | | |
|---|---|---|---|---|---|---|
| 30 | S-0521 | Negative for disease-causing or likely disease-causing variants | | | | |
| 31 | S-0627 | Negative for disease-causing or likely disease-causing variants | | | | |
| 32 | S-0723 | Negative for disease-causing or likely disease-causing variants | | | | |
| 33 | S-0998 | Negative for disease-causing or likely disease-causing variants | | | | |
| 34 | S-1049 | Negative for disease-causing or likely disease-causing variants | | | | |
| 35 | S-1121 | Negative for disease-causing or likely disease-causing variants | | | | |
| 36 | S-1368 | Negative for disease-causing or likely disease-causing variants | | | | |
| 37 | S-1473 | Negative for disease-causing or likely disease-causing variants | | | | |
| 38 | S-1477 | Negative for disease-causing or likely disease-causing variants | | | | |
| 39 | S-1482 | *GBA* | chr1:155204794G>A<br>c.1603C>T<br>p.Arg535Cys | Heterozygous | Pathogenic | Dominant/Recessive |
| | | *KANSL1* | chr17:44110478G>T<br>c.2805C>A<br>p.Thr935Thr | Heterozygous | Variant of Uncertain Significance (VUS) | Dominant |
| 40 | S-1483 | chr11p11.2 | chr11:(?_45889142)<br>_(47204298_?)del<br>(chr11p11.2 partial deletion) | Heterozygous | Pathogenic | Dominant |
| 41 | S-1487 | Negative for disease-causing or likely disease-causing variants | | | | |

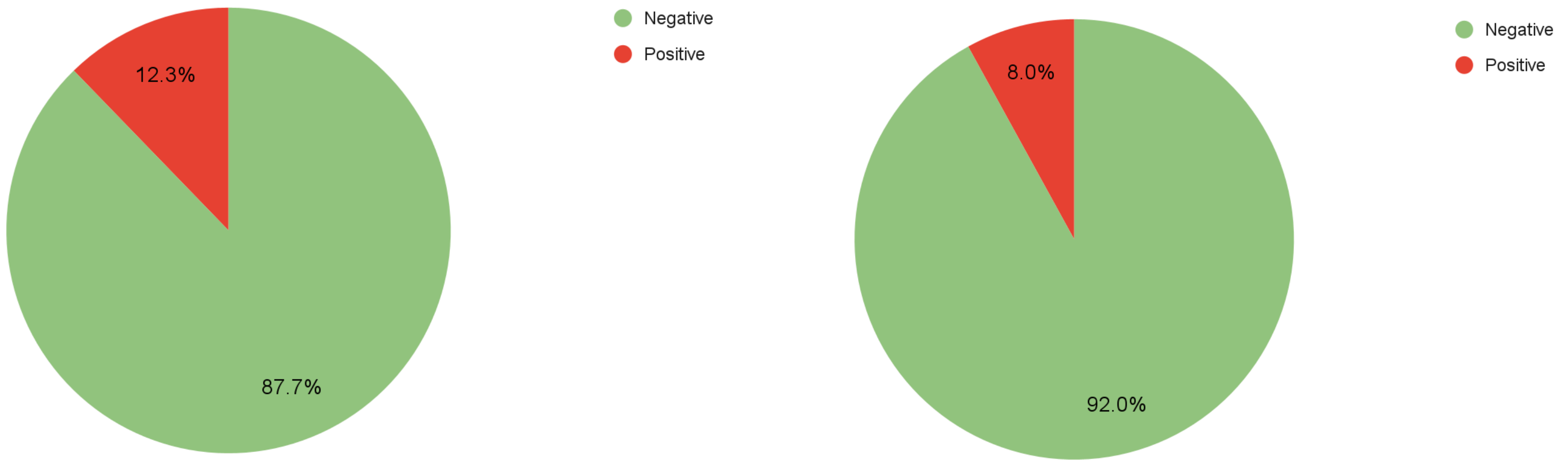


**Figure S1:** Pie charts represent the proportion of positive and negative cases among 136 samples analysed using low resolution workflow and 32 samples analysed using high resolution workflow. The detection rate was 12.3% for the low resolution group and 8.0% for the high resolution group.

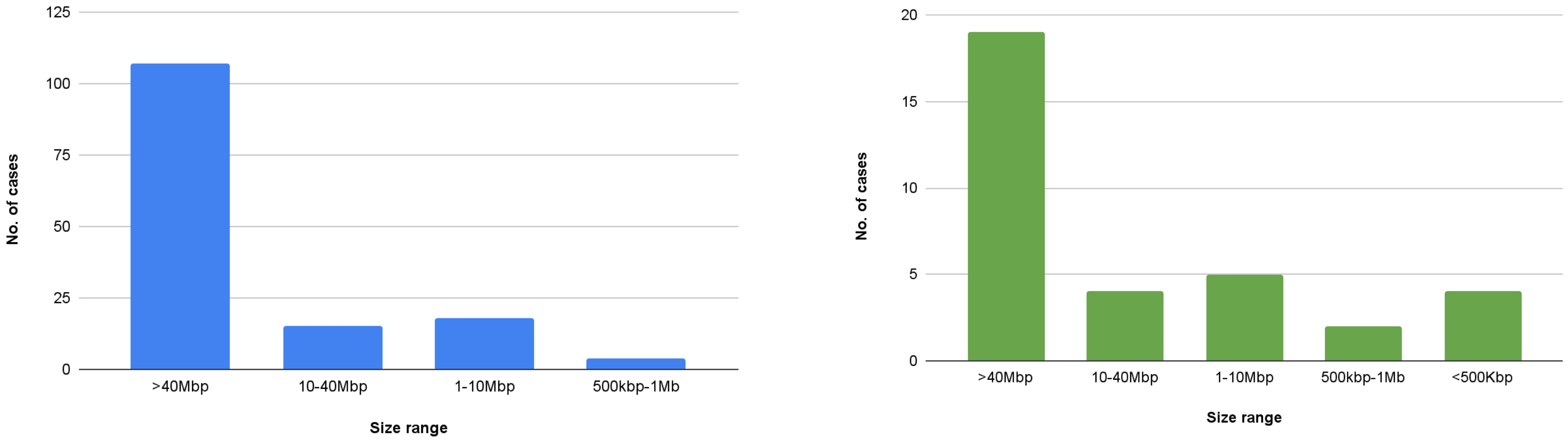


**Figure S2:** The bar graph represents the number of CNVs categorized by size in low-resolution (n = 147) and high-resolution (n = 31) cases. A. In low-resolution cases, the majority of CNVs (n = 107) were larger than 40 Mb, with fewer CNVs in the 10–40 Mb (n = 15), 1–10 Mb (n = 18), and 500 kb–1 Mb (n = 4) ranges. B. In high-resolution cases, most CNVs (n = 19) were also larger than 40 Mb, followed by 10–40 Mb (n = 4), 1–10 Mb (n = 5), 500 kb–1 Mb (n = 2), and <500 kb (n = 4). Notably, CNVs smaller than 500 kb were only detected in high-resolution cases.
